# Benchmarking Large Language Models for Extraction of International Classification of Diseases Codes from Clinical Documentation

**DOI:** 10.1101/2024.04.29.24306573

**Authors:** Ashley Simmons, Kullaya Takkavatakarn, Megan McDougal, Brian Dilcher, Jami Pincavitch, Lukas Meadows, Justin Kauffman, Eyal Klang, Rebecca Wig, Gordon Smith, Ali Soroush, Robert Freeman, Donald J Apakama, Alexander W Charney, Roopa Kohli-Seth, Girish N Nadkarni, Ankit Sakhuja

## Abstract

**Background:** Healthcare reimbursement and coding is dependent on accurate extraction of International Classification of Diseases-tenth revision – clinical modification (ICD-10-CM) codes from clinical documentation. Attempts to automate this task have had limited success. This study aimed to evaluate the performance of large language models (LLMs) in extracting ICD-10-CM codes from unstructured inpatient notes and benchmark them against human coder.

**Methods:** This study compared performance of GPT-3.5, GPT4, Claude 2.1, Claude 3, Gemini Advanced, and Llama 2-70b in extracting ICD-10-CM codes from unstructured inpatient notes against a human coder. We presented deidentified inpatient notes from American Health Information Management Association Vlab authentic patient cases to LLMs and human coder for extraction of ICD-10-CM codes. We used a standard prompt for extracting ICD-10-CM codes. The human coder analyzed the same notes using 3M Encoder, adhering to the 2022-ICD-10-CM Coding Guidelines.

**Results:** In this study, we analyzed 50 inpatient notes, comprising of 23 history and physicals and 27 progress notes. The human coder identified 165 unique codes with a median of 4 codes per note. The LLMs extracted varying numbers of median codes per note: GPT 3.5: 7, GPT4: 6, Claude 2.1: 6, Claude 3: 8, Gemini Advanced: 5, and Llama 2-70b:11. GPT 4 had the best performance though the agreement with human coder was poor at 15.2% for overall extraction of ICD-10-CM codes and 26.4% for extraction of category ICD-10-CM codes.

**Conclusion:** Current LLMs have poor performance in extraction of ICD-10-CM codes from inpatient notes when compared against a human coder.

## INTRODUCTION

Medical coding is an important part of the United States Healthcare System in the 21^st^ century. Healthcare organizations hire and train a substantial workforce proficient in abstracting medical codes from clinical records.^1^ This workforce then supports and submits claims for reimbursement adherent to regulatory requirements, handle insurance denials, support research endeavors, aid public health surveillance, and ensure a faithful representation of a patient’s medical history in the electronic health records (EHR).^2,3^ International Classification of Diseases (ICD) codes developed by the World Health Organization (WHO), are used to document specific diagnostic and procedural information such as medical history, surgical history and problem lists. The ICD codes are currently in their tenth revision (ICD-10). ^3^ ICD-10-Clinical Modification (ICD-10-CM) is a variant of ICD-10 adopted by the United States Government to add additional detail to the ICD-10 codes developed by WHO with around 68,000 diagnosis codes.^3^

Computerized assistive coding (CAC) technologies are currently used to improve the workflow of medical coding professionals. The American Health Information Management Association (AHIMA) defines CACs as “computer software that automatically generates a set of medical codes for review, validation, and use based upon provider clinical documentation.”^4^ Their performance, however, is still far below that of medical coding professionals.^5,6^ These CACs are thus used as semi-automated processes that augment human workflows.^4^

Recent studies have also explored the use of CACs powered with natural language processing but have been found wanting in handling of heterogenous, complex, and ambiguous medical terminology. For example, they struggle with common syntax such as references to instructions for patients to return if certain symptoms occur. The system does not recognize this as a hypothetical situation and will code those mentioned symptoms as if the patient is currently experiencing them.^7^

With the advent of large language models (LLM), there are new opportunities for further refinement of CACs. Our recent work has shown that current LLMs underperform in generating ICD codes when provided with a code description.^8^ With potential for applications of LLMs for billing in healthcare, in this study, we sought to benchmark current LLMs for extraction of ICD-CM codes from patient charts against a human coder.

## METHODS

For this study we evaluated multiple commercially available LLMs, including GPT 3.5, GPT 4, Claude 2.1, Claude 3, Gemini Advanced, and Llama 2-70b. With permission from American Health Information Management Association (AHIMA), we used deidentified patient notes from the AHIMA Vlab^9^ authentic patient cases for this study. AHIMA VLab is a virtual practice environment for health information education. It includes deidentified authentic patient charts that are used by students for coding exercises, chart analysis, general orientation to medical record forms and indexing.^9^ The AHIMA inpatient authentic patient cases are comprised of deidentified patient encounters; however, for our study we used inpatient notes that include a combination of both history-physical and progress notes. The AHIMA authentic patient cases can be accessed through My AHIMA Learning Center, an online portal^9^ at https://myahima.brightspace.com/d2l/home/6681.

These notes were presented to both the LLM’s and the human coder (AS) for extraction of ICD-10-CM codes. The human coder extracted ICD-10-CM codes in for billing purposes as current standard of practice. In addition to mastery level certification, the coder has 11 years of practical experience in medical coding and serves as an Assistant Professor for undergraduate students in a Health Informatics and Information Management Program, specializing in medical coding. We assigned each patient note a random number using ‘random’ module from Python 3.8.3. This was done to ensure blinding of the notes from the human coder. We used a standardized prompt for this study -“Please code the following note using the ICD-10 CM inpatient guidelines from 2022”. The human coder separately analyzed the same notes and extracted ICD-10-CM code(s) using 3M Encoder^10^, as is standard practice. Applicable 2022 ICD-10-CM Official Coding Guidelines for each note were applied by referring to the 2022 ICD-10-CM coding guidelines^3^ specific to inpatient settings. Each note was evaluated individually to ensure that the assigned codes adhere to the official coding guidelines.

### Statistical Analysis

We used the proportion of agreement to estimate the agreement between ICD-10-CM codes generated by LLMs and a human coder. This proportion is calculated by dividing the number of identical ICD-10-CM codes by the total number of ICD-10-CM codes identified by LLMs or the human coder for each case. We also calculated Cohen’s kappa to evaluate the agreement between LLMs and human coder. The Cohen’s kappa indicates a numeric rating of the degree of agreement between two raters, considering the degree of agreement that would be expected by chance^11^. We then evaluated the diagnostic performance of LLMs, compared to a human coder using precision and recall. We further did an exploratory analysis on a randomly chosen subset of 10% of patient notes to identify reasons for discrepancy between the human coder and LLMs in extraction of ICD-10-CM codes. For this analysis another human coder (MM) reviewed codes extracted by the LLMs in the chosen subset and identified reasons for discrepancy with the codes extracted by the human coder. In addition, we also assessed the performance of LLMs in extraction of Category ICD-10-CM codes. These are the 3-digit ICD-10-CM codes that identify the general categories of diagnoses. For example, N17 is the category ICD-10-CM code for Acute Kidney Injury which has further specific ICD-10-CM codes under it. We conducted all analyses using R version 4.2.2.^12^

## RESULTS

We included 50 patient notes in this study. This included 23 history and physicals, and 27 progress notes. The human coder extracted a total of 165 unique ICD-10-CM codes. As shown in **Figure 1A** the number of unique ICD-10-CM codes extracted by LLMs varied from 221 for Gemini Advanced to 658 for Llama 2-70b. The median [IQR] number of ICD-10-CM codes extracted by the human coder was 4 [2-6]. Among the LLMs, the number of ICD-10-CM codes extracted were as follows: GPT3.5: 7 [4-10], GPT4: 6 [4-8], Claude2.1: 6 [4-8], Claude3: 8 [6-10], Gemini Advanced: 5 [5-7], and Llama 2-70b: 11 [7-21]

**Figure 1.**
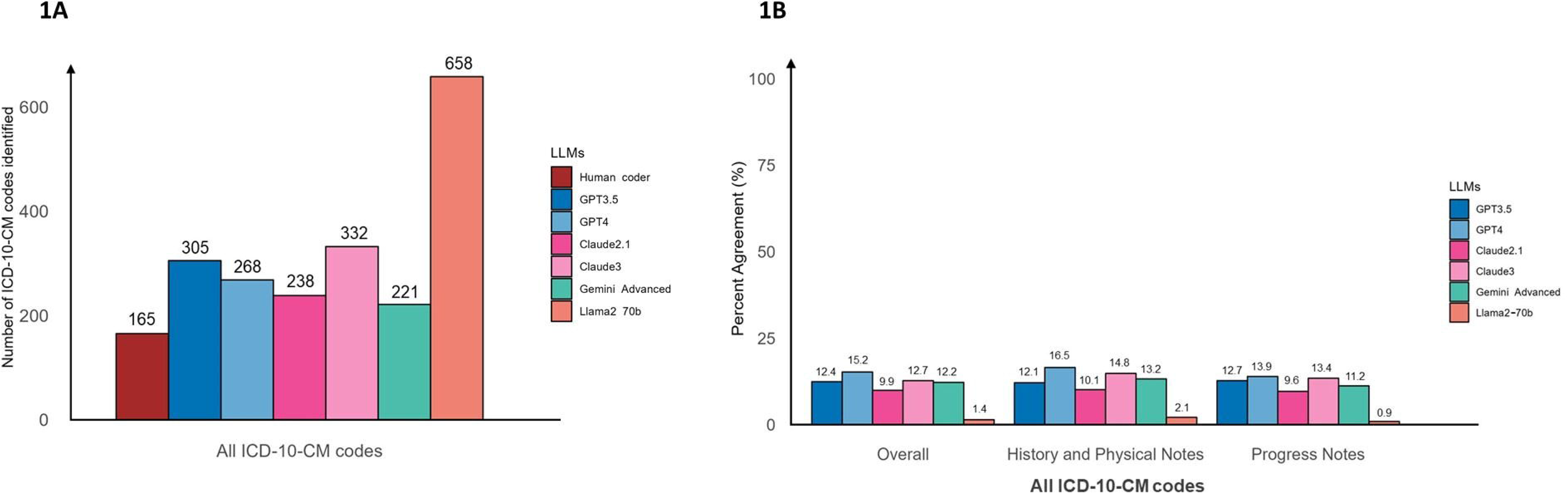
Number of ICD-10-CM codes identified (1A). Percentage agreement between individual LLMs and human coder in extraction of ICD-10-CM codes (1B)

### Performance

GPT4 achieved the highest percent agreement for ICD-10 code extraction among the LLMs and the human coder at 15.2%, followed by Claude3 (12.7%), GPT3.5 (12.4%), Gemini Advanced (12.2%), Claude2.1 (9.9%), and Llama 2-70b (1.4%) **(Figure 1B).** The Cohen’s kappa values were poor, ranging between −0.02 to 0.01, suggesting minimal to no agreement among LLMs when compared to the human coder **(Table 1)**. The reasons for discrepancy for the ICD-10-CM codes extracted by LLMs to that extracted by the human coder are shown in Table 2. Subgroup analysis of history and physical notes, as well as progress notes, revealed consistent results in percent agreement between LLMs and the human coder **(Figure 1B and Table 1).** When focusing solely on the primary diagnosis, Claude3 yielded a percent agreement of 26% and a kappa value of 0.25, followed by Claude2.1 (percent agreement 20% and kappa 0.20) and GPT4 (percent agreement 18% and kappa 0.17), respectively **(Supplementary Table 1).**

**Table 1.** Performance of LLMs for the ICD-10-CM code extraction compared to certified coding specialist.

|  | All notes | History and Physical notes | Progress notes |
| --- | --- | --- | --- |
| **GPT3.5** |  |  |  |
| **Kappa** | 0.01 | 0.02 | -0.01 |
| **Precision** | 0.17 | 0.18 | 0.16 |
| **Recall** | 0.31 | 0.30 | 0.31 |
| **GPT4** |  |  |  |
| **Kappa** | 0.01 | 0.02 | -0.01 |
| **Precision** | 0.21 | 0.23 | 0.19 |
| **Recall** | 0.35 | 0.37 | 0.33 |
| **Claude2.1** |  |  |  |
| **Kappa** | -0.02 | -0.02 | -0.01 |
| **Precision** | 0.15 | 0.16 | 0.14 |
| **Recall** | 0.22 | 0.21 | 0.24 |

| **Claude3** |  |  |  |
| --- | --- | --- | --- |
| **Kappa** | -0.01 | 0.01 | -0.02 |
| **Precision** | 0.16 | 0.19 | 0.14 |
| **Recall** | 0.36 | 0.40 | 0.32 |
| **Gemini Advanced** |  |  |  |
| **Kappa** | -0.02 | -0.02 | 0.01 |
| **Precision** | 0.19 | 0.21 | 0.18 |
| **Recall** | 0.25 | 0.27 | 0.24 |
| **Llama 2-70b** |  |  |  |
| **Kappa** | -0.02 | 0.01 | -0.02 |
| **Precision** | 0.02 | 0.03 | 0.01 |
| **Recall** | 0.07 | 0.08 | 0.06 |

**Table 2:** Reasons for discrepancy in extracted ICD-10-CM codes between individual LLMs when compared again the human coder (evaluated in 10% random subset)

| **Issue** | **GPT**<br>**3.5** | **GPT 4** | **Claude**<br>**2.1** | **Claude**<br>**3** | **Gemini**<br>**Advanced** | **Llama**<br>**2-70b** |
| --- | --- | --- | --- | --- | --- | --- |
| **Inappropriate coding for Symptoms, Signs of Conditions** | 4<br>(20%) | 1 (3%) | 5 (22%) | 4 (12%) | 1 (4%) | 10<br>(16%) |
| **Coding for diagnoses not confirmed by providers** | 7<br>(35%) | 17<br>(60%) | 6 (26%) | 19<br>(57%) | 5 (20%) | 34<br>(54%) |
| **Use of non-specific codes** | 5<br>(25%) | 1 (3%) | 1 (4%) | 3 (10%) | 2 (8%) | 12<br>(19%) |
| **Hallucinations** | 2<br>(10%) | 6<br>(21%) | 8 (35%) | 6 (18%) | 7 (28%) | 6<br>(10%) |
| **Missed Diagnosis** | 2<br>(10%) | 3<br>(10%) | 3 (13%) | 1 (3%) | 10 (40%) | 1 (1%) |
| **Total discrepant codes** | 20 | 28 | 23 | 33 | 25 | 63 |

### Category ICD-10-CM codes

There were 146 unique category ICD-10-CM codes extracted by the human certified code**r (Figure 2A)**. GPT4 achieved the highest percent agreement at 26.4%, followed by GPT3.5 (23.6%), Claude3 (21.3%), Claude2.1 (20.8%), Gemini Advanced (20.6%), and Llama 2-70b (10%), respectively **(Figure 2B).** The Cohen’s kappa values were again poor, ranging between −0.01 to 0.03, suggesting minimal to no agreement among LLMs when compared to human coder **(Table 3)**. When focusing on the primary diagnosis, Claude2.1 and Claude3 achieved the best performance with a percent agreement of 36% and a kappa value of 0.35, followed by GPT4 (percent agreement 34%, kappa 0.33), and Gemini Advanced (percent agreement 30%, kappa 0.31) (Supplementary Table 2).

**Figure 2.**
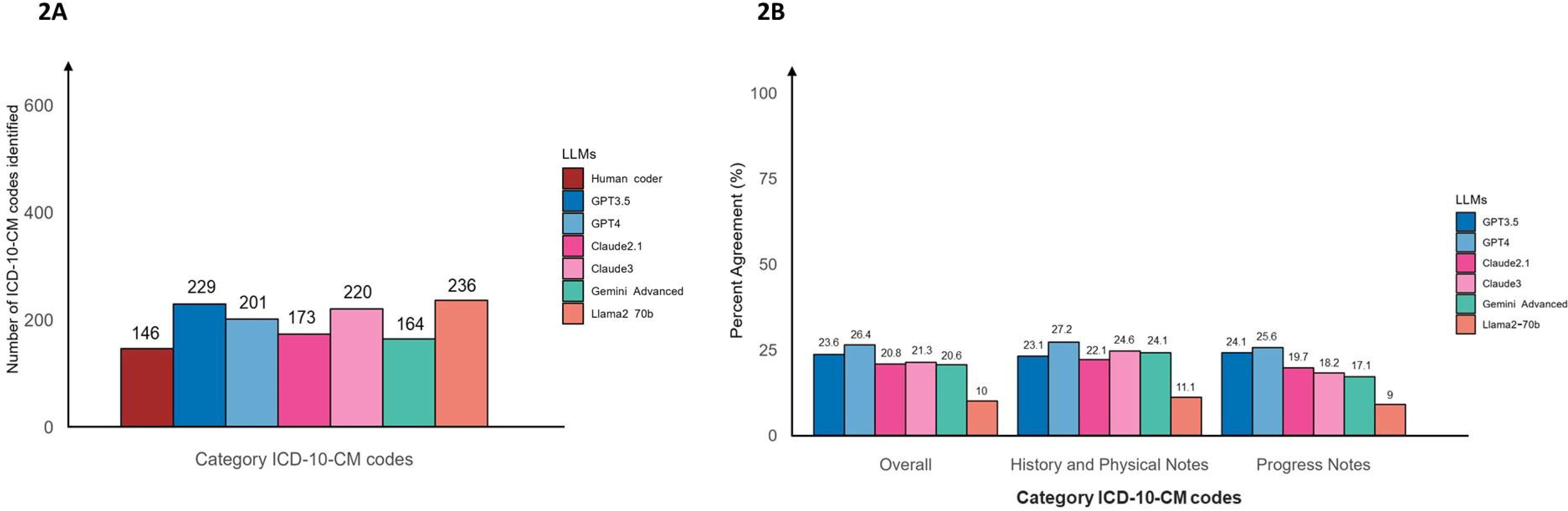
Number of category ICD-10-CM codes identified (2A). Percentage agreement between individual LLMs and human coder in extraction of category ICD-10-CM codes (2B)

**Table 3.** Performance of LLMs for the category ICD-10-CM code extraction compared to certified coding specialist.

|  | **All notes** | **History and Physical notes** | **Progress notes** |
| --- | --- | --- | --- |
| **GPT3.5** |  |  |  |
| **Kappa** | 0.02 | 0.01 | -0.01 |
| **Precision** | 0.31 | 0.29 | 0.32 |
| **Recall** | 0.51 | 0.52 | 0.5 |
| **GPT4** |  |  |  |
| **Kappa** | 0.03 | 0.01 | 0.02 |
| **Precision** | 0.34 | 0.35 | 0.33 |
| **Recall** | 0.55 | 0.56 | 0.55 |
| **Claude2.1** |  |  |  |
| **Kappa** | -0.01 | 0.01 | -0.02 |
| **Precision** | 0.29 | 0.32 | 0.26 |
| **Recall** | 0.43 | 0.41 | 0.44 |
| **Claude3** |  |  |  |
| **Kappa** | -0.01 | -0.01 | 0.01 |
| **Precision** | 0.26 | 0.30 | 0.22 |
| **Recall** | 0.53 | 0.58 | 0.49 |
| **Gemini Advanced** |  |  |  |
| **Kappa** | -0.01 | -0.01 | 0.02 |
| **Precision** | 0.31 | 0.35 | 0.26 |
| **Recall** | 0.39 | 0.44 | 0.34 |
| **Llama 2-70b** |  |  |  |
| **Kappa** | -0.01 | 0.01 | -0.01 |
| **Precision** | 0.13 | 0.15 | 0.12 |
| **Recall** | 0.29 | 0.30 | 0.28 |

## DISCUSSION

In this study we have benchmarked the performance of LLMs in extracting ICD-10-CM codes from narrative documentation in patient charts. We conducted this evaluation using a comparative analysis on the performance of these models against that of a human coder. The LLMs evaluated in this study included GPT-3.5, GPT 4, Claude 2.1, Claude 3, Gemini Advanced, and Llama 2-70b. We found that all evaluated LLMs had poor concordance in extraction of ICD-10-CM codes when compared to a human coder. GPT 4, however, achieved the best performance in both, overall extraction of ICD-10-CM codes and category ICD-10-CM codes. When focusing only on primary diagnosis, Claude 3 showed the best performance across extraction of both overall ICD-10-CM codes and category ICD-10-CM codes. We found similar results with extraction of both, entire ICD-10-CM code and just category ICD-10-CM codes. We have further evaluated the reasons for discrepancy in extraction of ICD-10-CM codes by individual LLMs when compared against the human coder.

Since the introduction of GPT 3.5, there has been a steady interest in exploring the capabilities of LLMs in various areas. Recent studies have shown that GPT 3.5, one of the first LLM models available, achieved a passing score in the United States Medical Licensure Exam (USMLE)^13^ and passed two portions of the Bar Exam – evidence and torts.^14^ These are complex examinations that are specific to their professional fields – USMLE for medicine and bar exam for law. USMLE questions span a diverse range of topics in medicine that include clinical medicine, basic science and bioethics. Similarly, passing a Bar Exam requires an in depth understanding of the law and the legal language. The fact that an LLM that has not been trained specifically for this purpose, can perform so well in such specific professional examinations has led to a lot of excitement about the potential of such models.

The LLMs, however, fail to replicate similar performance for more specific tasks. For example, in a study that evaluated the ability of GPT 3.5 to answer questions related to the field of nephrology, the results were much less impressive with only 51% accuracy rate^15^. The authors used questions from Nephrology Self-Assessment Program and Kidney Self-Assessment Program.^16,17^ Both of these resources are used to enhance and refresh clinical knowledge in the field of nephrology and for preparation of the American Board of Internal Medicine Nephrology Board Examination. This was way below the passing threshold of 75% for Nephrology Self-Assessment Program and 76% for Kidney Self-Assessment Program. Another recent study that evaluated the performance of LLMs on Nephrology Self-Assessment Program and Kidney Self-Assessment Program found that GPT 4 achieved a much better performance with 73.3% correct answers^18^, still however, below the passing threshold. Performance of Claude 2 and Llama was much worse with only 54.4% and 30.6% correct responses, respectively. Another study that evaluated GPT 3.5’s performance on questions from similar resources but with a focus on glomerular diseases, a group of highly specific kidney diseases, found that GPT 3.5’s accuracy further dropped down to 45%^15^. LLMs have shown similar suboptimal performance in self-assessment tests designed for other specialties such as gastroenterology^19^, ophthalmology^20^ and urology^21^. Thus, it seems that even though LLMs may perform well with general professional examinations, they do not perform well when more specific knowledge of the field is required.

It is therefore not surprising that LLMs in our study were unable to perform well in the highly specialized task of extracting ICD-10-CM codes from inpatient notes. The training required to become a medical coder is complex and includes a comprehensive education in medical terminology, pathophysiology, anatomy, and pharmacology, in addition to the coding terminology itself. The coders must learn to parse through the medical records and tease out the right diagnostic codes, while separating out the verbiage that discusses symptomatology or warning signs. It therefore requires an in-depth understanding of ICD-10-CM system, clinical documentation, and a great command of English language.

Our study highlights the limitations of LLMs while extracting ICD-10 CM codes from inpatient notes. While the human coder extracted total 165 unique ICD-10-CM codes, the total of unique ICD-10-CM codes extracted by the LLMs were much higher. Gemini Advanced extracted 221 ICD-10-CM codes - the least amount among the LLMs studied. Claude 2.1 was next and extracted 238 ICD-10-CM codes. This was followed by 268 ICD-10-CM codes with GPT4, 305 with GPT-3.5, 332 with Claude3 and finally 658 ICD-10-CM codes with Llama 2-70b – the highest number of them all. As shown in Table 2, there were multiple reasons for this discrepancy. For example, some of these codes resulted from the inability of individual LLMs to distinguish symptom codes from diagnosis codes as established in the ICD-10-CM Official Coding Guidelines. According to guidelines, conditions and signs or symptoms codes falling within categories R00-R94 should only be used when more specific diagnosis cannot be made even after all the facts bearing on the case have been investigated, and in cases in which a more precise diagnosis was not available for any other reason.^3^ For example, in a case where the patient presented with chest pain, cough and fatigue but was diagnosed with upper respiratory infection, the codes for chest pain, cough, fatigue were also extracted by one of the LLMs. Because there was a precise diagnosis code for the upper respiratory infection, the sign and symptom codes were not necessary, therefore leading to an inflated code count.

The LLMs at times also failed to accurately identify all the secondary diagnoses for those cases or assigned additional diagnoses without available supporting clinical documentation. In one notable instance, the LLM identified elevated sodium levels listed within the lab results and assigned the diagnosis code E87.0 (hyperosmolality and hypernatremia) without any corresponding physician documentation to validate the diagnosis. This is an example of the LLM disregarding the coding guidelines outlined in Section I. A. 19, which emphasizes that diagnosis codes should be assigned solely based on the diagnostic statements provided by the healthcare provider within the notes, rather than relying on the clinical criteria used by the provider to establish the diagnosis (i.e. lab values).^3^ As shown in table 2, there were also instances of hallucinations where LLM coded diagnoses not present anywhere in the note, and use of non-specific codes.

The identified trend in the LLM code assignments sequencing further suggests that the systems arranged the codes based on numerical order as abstracted directly from the clinical notes provided, rather than prioritizing the codes based on hierarchical coding guidance. This also emphasizes the limitations in LLMs understanding of the hierarchy involved in coding sequencing.

Our results are consistent with prior literature showing mediocre performance of LLMs when working with ICD codes. Spark NLP, a much smaller NLP model, has shown much better performance in extraction of ICD-10-CM codes in comparison to GPT 3.5 and GPT 4.^22^ In comparison to a success rate of 76% achieved by Spark NLP, the overall accuracies of GPT 3.5 and GPT 4 were only 26% and 36%, respectively. Recent literature has shown that LLMs struggle to generate diagnosis when provided with ICD codes.^23^ Our recent work has further shown that LLMs have difficulty in generating billing codes when providing code descriptions.^8^ Among GPT 3.5, GPT 4, Gemini Advanced and Llama 2-70b, we found that GPT 4 had the best performance to generate ICD-10-CM codes when provided with code descriptions. The performance will still poor at only 33.9% match rate. We found similar results in this study where GPT 4 had the best, albeit still poor performance in extraction of ICD-10-CM codes when compared against a human coder. Our current work builds systematically on the evolving LLM literature and benchmarks their performance against that of a human coder. As human coders are used by hospitals for extraction of ICD codes as current standard of practice, this study provides an effective benchmark for future LLM research in this highly specialized area.

Though our study provides important insights into the performance of LLMs for extraction of ICD-10-CM codes, it is important to interpret these results while understanding the limitations of the study. We only investigated extraction of ICD-10-CM codes based on inpatient notes and the results are therefore not generalizable to extraction of ICD-10-CM codes based on outpatient notes or to extraction of ICD-10 procedure codes. As our goal was to benchmark the performance of LLMs to that of a human coder, we used a standardized prompt to generate responses. It is important to acknowledge that utilization of different prompts can elicit differing responses. Our study also does not utilize retrieval augmented generation, which could potentially further enhance the performance of LLMs.

In summary, our study benchmarks the performance of LLMs in the highly specialized task of extraction of ICD-10-CM codes from inpatient notes, against a human coder. Although GPT 4 exhibited the highest overall performance in ICD-10-CM code extraction, it still fell short. Future investigations should focus on advanced prompt engineering, incorporating retrieval augmented generation and fine-tuning models to enhance the performance of LLMs ICD-10-CM extraction.

## Data Availability

All data produced are available online at https://myahima.brightspace.com/d2l/home/6681

## Acknowledgements

AHIMA VLab for permission to use the deidentified inpatient notes for the study.

## Funding

This study was supported by National Institutes of Health (NIH) grant K08DK131286 (AS)

## Competing Interests

GNN is a founder of Renalytix, Pensieve, Verici and provides consultancy services to AstraZeneca, Reata, Renalytix, Siemens Healthineer and Variant Bio, serves a scientific advisory board member for Renalytix and Pensieve. He also has equity in Renalytix, Pensieve and Verici. All remaining authors have declared no conflicts of interest.

## Notes

### Funding Statement

This work was supported by the National Institutes of Health (NIH) grants K08DK131286 awarded to Ankit Sakhuja. The content is solely the responsibility of the authors and does not necessarily represent the official views of the National Institutes of Health.

### Summary of Updates

Funding statement updated to ensure it can be assigned PMCID number

